# Leveraging polygenic enrichments of gene features to predict genes underlying complex traits and diseases

**DOI:** 10.1101/2020.09.08.20190561

**Authors:** Elle M. Weeks, Jacob C. Ulirsch, Nathan Y. Cheng, Brian L. Trippe, Rebecca S. Fine, Jenkai Miao, Tejal A. Patwardhan, Masahiro Kanai, Joseph Nasser, Charles P. Fulco, Katherine C. Tashman, Francois Aguet, Taibo Li, Jose Ordovas-Montanes, Christopher S. Smillie, Moshe Biton, Alex K. Shalek, Ashwin N. Ananthakrishnan, Ramnik J. Xavier, Aviv Regev, Rajat M. Gupta, Kasper Lage, Kristin G. Ardlie, Joel N. Hirschhorn, Eric S. Lander, Jesse M. Engreitz, Hilary K. Finucane

## Abstract

Genome-wide association studies (GWAS) are a valuable tool for understanding the biology of complex traits, but the associations found rarely point directly to causal genes. Here, we introduce a new method to identify the causal genes by integrating GWAS summary statistics with gene expression, biological pathway, and predicted protein-protein interaction data. We further propose an approach that effectively leverages both polygenic and locus-specific genetic signals by combining results across multiple gene prioritization methods, increasing confidence in prioritized genes. Using a large set of gold standard genes to evaluate our approach, we prioritize 8,402 unique gene-trait pairs with greater than 75% estimated precision across 113 complex traits and diseases, including known genes such as *SORT1* for LDL cholesterol, *SMIM1* for red blood cell count, and *DRD2* for schizophrenia, as well as novel genes such as *TTC39B* for cholelithiasis. Our results demonstrate that a polygenic approach is a powerful tool for gene prioritization and, in combination with locus-specific signal, improves upon existing methods.

## INTRODUCTION

Genome-wide association studies (GWAS) have identified thousands of genetic loci associated with common complex traits and diseases^1^. Nonetheless, for the vast majority of GWAS significant loci, the identity of the causal gene(s) underlying the association remains unknown, limiting the biological insight gained into common disease mechanisms^2,3^. There are several major challenges to pinpointing the causal gene. First, linkage disequilibrium (LD) between variants masks the identity of the causal variant^4^. Second, most associated loci do not contain coding variants but, instead, the causal variant acts through gene regulatory mechanisms^3^, and incomplete maps from regulatory element to gene hinder causal gene identification^5^. Many computational approaches try to resolve these challenges^6–10^, yet methods in the field of gene prioritization often fail to nominate causal genes with high confidence.

Gene prioritization strategies can be placed into two broad categories: first, locus-based methods that leverage local GWAS data by connecting the causal variants to the causal gene(s) using protein coding variants, genomic distance, enhancer-gene maps^11–16^, or eQTLs^7,8^; second, similarity-based methods that search for global patterns in associated genes and nominate those with similar functions, pathways, or network connections^6,10^. Across both categories, existing methods lack consensus and have high false positive rates^17^. At the same time, related work also suggests that combining results from different methods, specifically combining locus-based and similarity-based approaches, can yield better predictions^18^. Among similarity-based approaches, no method we are aware of leverages the full genome-wide association data, despite the fact that single-nucleotide polymorphisms (SNPs) that do not reach genome-wide significance explain the majority of narrow sense heritability for most complex traits^19^. Moreover, recently-generated single-cell RNA-seq datasets hold promise for more accurately characterizing shared functions among genes and thus improving the accuracy of similarity-based gene prioritization. Yet, to our knowledge, these data sets have not been systematically leveraged in this way.

Here, we propose a new similarity-based gene prioritization method, a gene-level Polygenic Priority Score (PoPS), that leverages the full polygenic signal, excluding GWAS data at the locus of interest, and incorporates data about genes from a variety of sources, including 73 publicly available single-cell RNA-seq data sets. PoPS is computationally efficient and requires only summary statistics and an LD reference panel. Across 113 complex traits and diseases, we show that PoPS outperforms other similarity-based gene prioritization methods. Using a unique set of gold standard genes, we show that, while neither PoPS nor any existing locus-based method based on non-coding variants alone achieves precision above 52%, PoPS combined with these locus-based gene prioritization methods achieves precision between 75% and 86% and prioritizes 8,395 unique gene-trait pairs. Finally, in several illustrative cases where the causal gene is known, we show that PoPS correctly identifies the correct gene even when the causal variant has been experimentally demonstrated to regulate multiple genes.

## RESULTS

### Overview of PoPS

Our method, PoPS, is predicated on the assumption that causal genes share functional characteristics. Specifically, we assume genes whose physical locations on the genome are near associated SNPs and who share similar biological annotations are most likely to be causal. PoPS uses gene-level associations computed from GWAS summary statistics to learn joint polygenic enrichments of gene features derived from cell-type specific gene expression, biological pathways, and protein-protein interactions (PPI). To nominate causal genes, PoPS then assigns a priority score to every protein coding gene according to these enrichments (**Fig. 1**).

First, PoPS applies MAGMA^9^ to compute gene-level association statistics and gene-gene correlations using GWAS summary statistics and LD information from an ancestry matched reference panel (see Methods). Next, PoPS performs marginal feature selection by using MAGMA to perform enrichment analysis for each gene feature separately. MAGMA tests a gene feature, *f*, for enrichment by modeling the gene-level associations, *y*, by

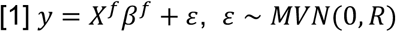

where *X*^*f*^ is a column vector corresponding to gene feature *f* (*e.g*. a binary indicator of membership in a pathway), and *R* is a covariance matrix designed to account for the LD between nearby genes computed from a reference panel. The model is fit by generalized least squares (GLS), and MAGMA reports both 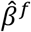 and a p-value for the hypothesis that *β*^*f*^ ≠ 0. We use these results to perform marginal feature selection, retaining only features that pass a nominal significance threshold (P < 0.05), to reduce the noise and computational complexity of fitting the joint model.

Second, PoPS computes a joint enrichment of all selected features simultaneously by replacing the vector *X*^*f*^ in Equation 1 with a matrix *X* that includes these features. (See **Supplementary Fig. 1** for comparison of model-fitting choices.) In the joint model, we additionally include a matrix *C* of gene-level covariates, (*e.g.* length of the gene, see Methods).

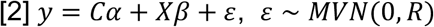

We extend the GLS method used by MAGMA to incorporate L_2_ regularization to account for the large number of features and improve test set prediction. We estimate 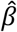 in a leave one chromosome out (LOCO) framework, obtaining estimates 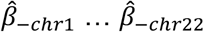.

Finally, PoPS computes polygenic priority scores for each gene, *g*, on a chromosome, *i*, by multiplying its row vector of gene features, *X*_*g*_, by 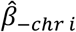. For example, to compute priority scores for gene *g* on chromosome 1, we compute 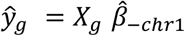. We refer to 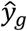 as the polygenic priority score (PoP score) for gene *g*. Ultimately, the PoP score for a gene is independent of the GWAS data on the chromosome where the gene is located. We say PoPS prioritizes a gene if it is in a 1 Mb locus centered on a genome-wide significant variant and has the highest PoP score of genes in the locus (see Methods).

**Figure 1.**
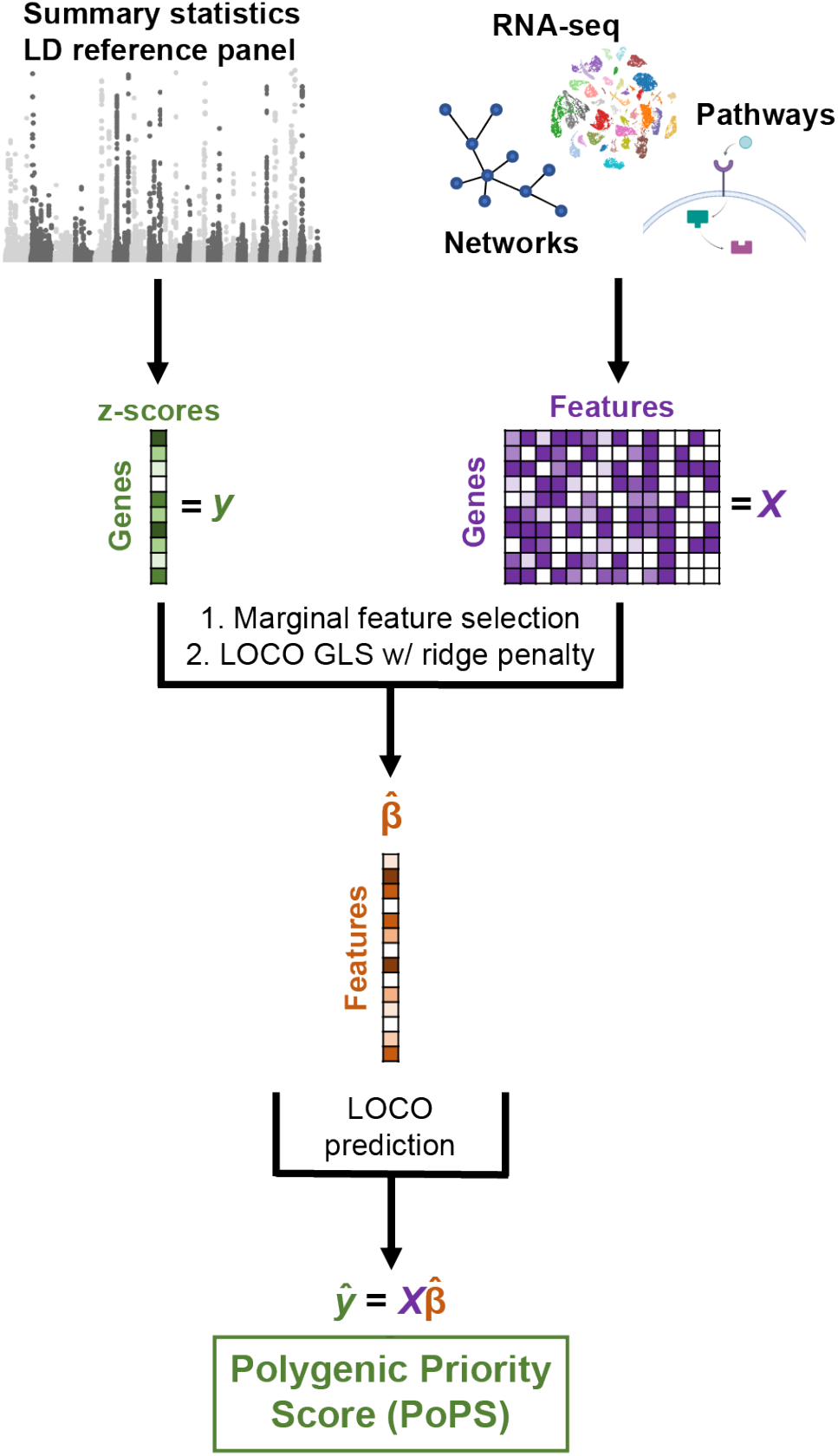
Overview of PoPS. We compute gene-level z-scores from GWAS summary statistics with an LD reference panel, using MAGMA. We create gene features from gene expression data, biological pathways, and predicted PPI networks and use marginal feature selection to narrow down to a smaller set of features most likely to be relevant. We then fit a linear model for the dependence of gene-level associations on gene features in a leave one chromosome out (LOCO) framework using generalized least squares (GLS) to account for LD and an L2 penalty to account for the large number of features. This results in a vector of joint polygenic enrichments of gene features, 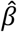, which we use to assign gene priority scores on the left-out chromosome.

### Application of PoPS to 113 complex traits

We applied PoPS to 18 diseases with publicly available GWAS summary statistics and 95 complex traits from the UK Biobank (**Supplementary Table 1**) using EUR individuals from the 1000 Genomes Project^20^ as a reference panel (see Methods). The full set of gene features used in these analyses included 57,543 total features – 40,546 derived from gene expression data, 8,718 extracted from a protein-protein interaction network^21^, and 8,479 based on pathway membership^22–25^ (**Supplementary Table 2**; see Methods). After marginal feature selection, 3,717 to 27,254 features per trait were included in the predictive model (**Supplementary Fig. 2**). For each trait, we score 18,383 protein coding genes and prioritize one gene in each genome-wide significant locus. In total, PoPS prioritized 18,179 unique gene-trait pairs in 25,341 loci across 113 complex traits.

### Evaluation of PoPS

To evaluate the performance of PoPS for prioritizing likely causal genes, we avoided benchmarking using curated sets of gold standard genes that may be biased towards well-studied genes or genes in well-characterized pathways. We instead evaluated PoPS with two metrics unaffected by prior knowledge of trait etiology by taking advantage of the fact that PoPS does not use GWAS data at the locus of interest when scoring genes. First, we applied the Benchmarker method^26^, which estimates the average contribution of SNPs near genes with high priority scores to per SNP heritability (see Methods). We refer to this quantity as normalized **τ**. After correction for multiple testing, we found our estimates for normalized **τ** were significantly greater than zero for 106 of 113 traits tested, indicating that genes with high PoP scores are enriched for heritability, even after accounting for the contributions of 53 other genomic annotations (**Fig. 2a, Supplementary Table 3**). As a second evaluation metric, we focused on the performance of PoPS in GWAS significant loci. Following the assumption that the causal gene is often the closest gene to the lead variant in the locus^18^, we tested whether PoPS prioritized genes were more often the closest gene than expected by chance (see Methods). Although this test is underpowered for traits with a small number of significant loci, we found that PoPS prioritized genes were significantly enriched for being the closest gene for 86 of 113 traits tested after Bonferroni correction (**Fig. 2b, Supplementary Table 3**). These results imply that, even though PoPS does not leverage any GWAS data at the locus, PoP scores are informative about the causal gene(s) at a locus.

### Comparison to other similarity-based methods

After evaluating PoPS on its own, we investigated how PoPS compares to existing similarity-based methods: DEPICT^6^ and NetWAS^10^. We applied PoPS, DEPICT, and NetWAS to the same set of 113 summary statistics and, again, used Benchmarker^26^ to evaluate the methods by comparing estimates of normalized **τ**. We found that PoPS showed the strongest performance among the three methods for all 113 traits tested (**Supplementary Table 3**). In a meta-analysis over 46 traits chosen to have low genetic correlation (see Methods), we confirmed that PoPS significantly outperformed both DEPICT and NetWAS overall (**Fig. 2c**). Similarly, we found that genes prioritized by PoPS showed the greatest enrichment for being the closest gene for 96 of 113 traits tested, and significantly outperformed DEPICT and NetWAS in a meta-analysis over independent traits (**Supplementary Fig. 3a, Supplementary Table 3**). We attribute the superior performance of PoPS to leveraging the full genome-wide association signal and integrating a broader array of gene features.

**Figure 2.**
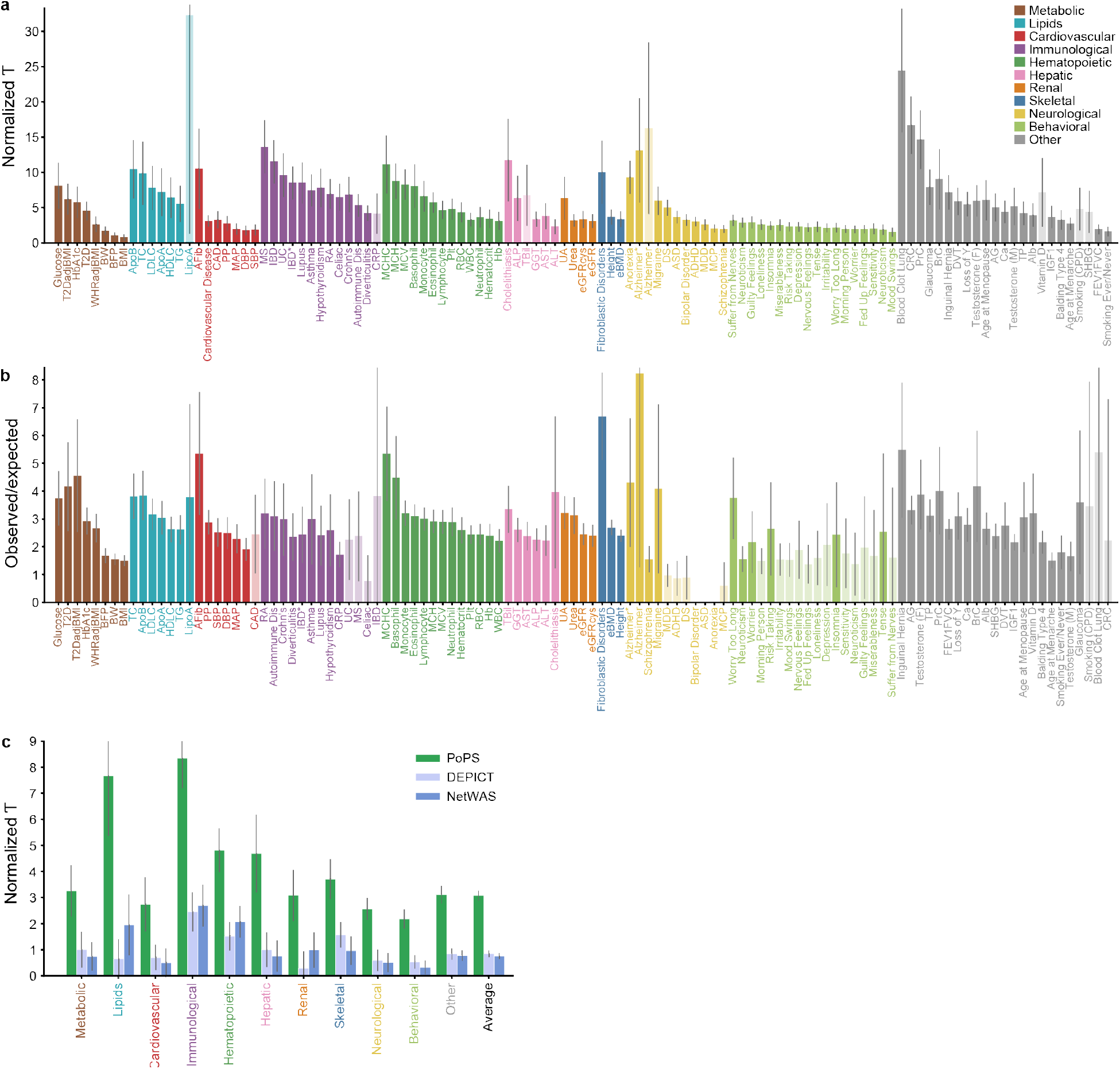
Evaluation of PoPS and comparison to other similarity-based methods. **a**, Results using Benchmarker to evaluate PoPS, grouped by trait domain and sorted by the lower bound of the 95% confidence interval of normalized **τ**. Normalized **τ** provides an estimate for the average contribution of SNPs near genes with high priority scores to per SNP heritability, normalized by average per SNP heritability. Error bars represent 95% confidence intervals. Dark bars passed the Bonferroni significance threshold (P < 0.05/113). For IBD and Alzheimer’s we retained summary statistics from both UK Biobank and other publicly available sources with a greater sample size; asterisk (*) denotes the public version. **b**, Results using closest gene enrichment to evaluate PoPS, grouped by trait domain and sorted by lower bound of the 95% confidence interval of the observed/expected ratio. Error bars represent 95% confidence intervals. P-values were computed using a normal approximation to the null distribution, and dark bars passed the Bonferroni significance threshold (P < 0.05/113). **c**, Results using Benchmarker to compare similarity-based gene prioritization methods, meta-analyzed within each trait domain across independent traits. Error bars represent 95% confidence intervals.

### Most informative gene features used by PoPS

We next evaluated the relevance of each category of features included in the PoPS model: gene expression, pathways, and PPI networks. We created three alternate versions of PoPS, training on features from each category separately, to produce three new sets of results for each phenotype. We applied Benchmarker to evaluate which set of features demonstrated the strongest performance on its own as measured by normalized **τ**. We meta-analyzed the estimates for normalized **τ** within 11 trait domains across 46 independent traits. Overall, we found that including all features yielded the strongest performance, followed by gene expression and pathways performing similarly, and finally followed by PPI networks (**Fig. 3a, Supplementary Table 3**). We similarly found that including all features showed the strongest enrichment using the closest gene metric (**Supplementary Fig. 3b, Supplementary Table 3**). We additionally evaluated the relevance of gene expression features based on bulk and single-cell RNA seq separately and found that the single-cell features performed significantly better than the bulk features by both the Benchmarker and closest gene metrics (**Supplementary Fig. 4, Supplementary Table 3**).

**Figure 3.**
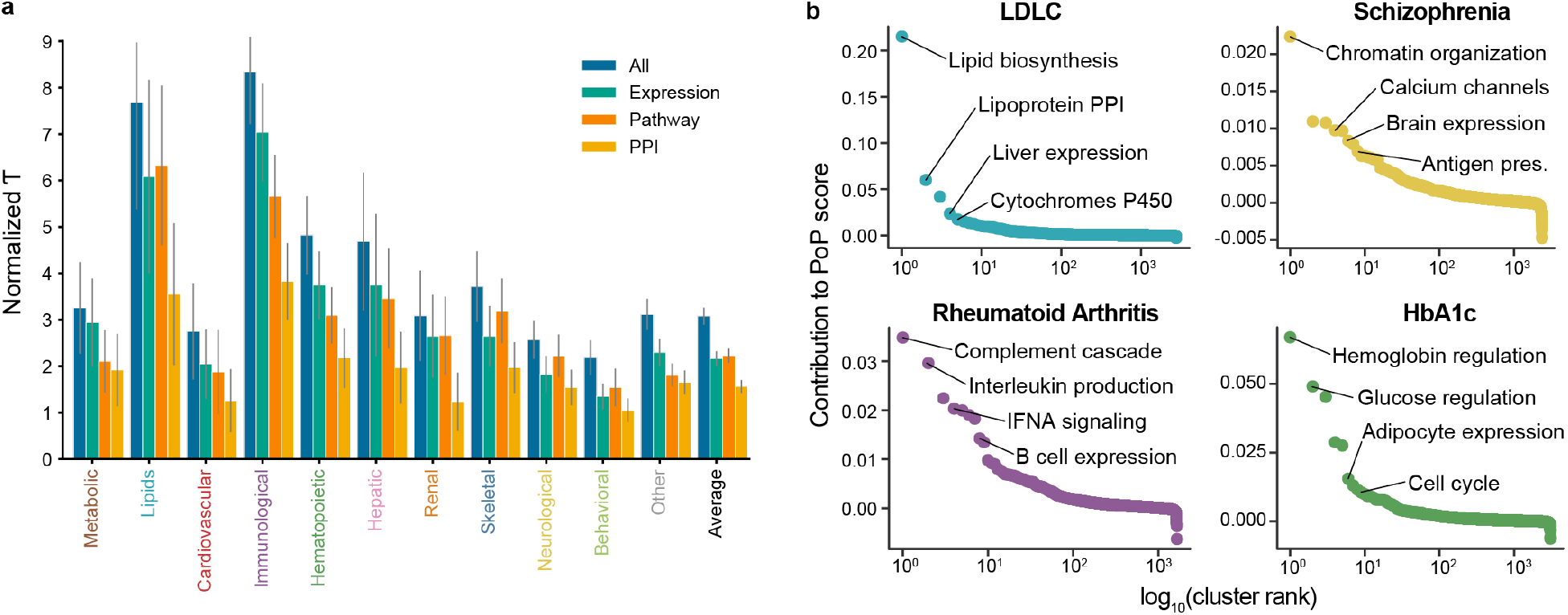
Most informative gene features used by PoPS. **a**, Results using Benchmarker to compare PoPS using different feature sets, meta-analyzed within each trait domain across independent traits. Error bars represent 95% confidence intervals. **b**, Rank-order plots for selected traits highlighting the feature clusters with the greatest contribution to the PoP scores of prioritized genes.

To better understand the relevant tissues, cell-types, and pathways learned by PoPS, we investigated which groups of features were most informative for prioritized genes. Because many highly correlated features were included in the joint model for PoPS, the individual coefficients, 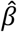, lacked direct interpretability. We instead grouped related features for a trait by performing hierarchical clustering on the selected features (see Methods). For each cluster, we computed the total contribution of the features in the cluster to the PoP scores of prioritized genes. We ranked clusters by their PoP score contribution and annotated top-ranked clusters (**Fig. 3b**). The features that composed the top clusters recapitulated known trait biology and included examples from each type of feature (**Supplementary Data 1**). For LDL cholesterol, we observed clusters composed of lipid synthesis pathways, lipoprotein PPI-interactions, and liver gene expression features^27^. For HbA1c, a test that measures average blood sugar levels but is also affected by red blood cell levels^28^, we observed both glucose and hemoglobin related clusters of features. For rheumatoid arthritis, an autoimmune disease^29^, we observed a range of immune features describing expression, signaling, and production of immune cells. Finally, for schizophrenia, we observed clusters corresponding to several distinct mechanisms previously implicated in schizophrenia including chromatin remodeling^30^ and calcium channel dysfunction^30,31^. Taken together, these results suggest that PoPS is able to prioritize the causal genes underlying complex traits and diseases by learning biologically relevant properties from multiple types of gene features.

### Comparison to locus-based methods

Noting previous work on the utility of ensemble gene prioritization^18^, we hypothesized that combining PoPS, a similarity-based method, with existing locus-based gene prioritization strategies would allow us to nominate genes with higher confidence than with either method alone – particularly for non-coding GWAS signals, where identifying the causal gene has been challenging. Towards that end, we applied several additional gene prediction methods to the set of 95 traits from the UK Biobank (see Methods), where we had not only summary statistics, but fine-mapping results (Ulirsch and Kanai, *In preparation)*. We focused our analysis on the 22,548/24,728 (91%) of the 95% credible sets (CSs) that did not contain a fine-mapped (posterior inclusion probability, PIP > 0.10) coding variant, and evaluated the following methods:

1. We overlapped fine-mapped (PIP > 0.1) non-coding variants with predicted enhancer-gene connections from (**a**) correlating enhancer and promoter activity (E-P correlation)^15,16^, (**b**) 3-D loops from promoter capture Hi-C (PCHi-C)^12–14^, and (**c**) activity-by-contact (ABC)^11^ maps to identify genes regulated by fine-mapped variants.
2. We incorporated eQTL data and (**a**) applied TWAS^8^ with GTEx v7^32^ weights to identify significantly associated genes and (**b**) computed co-localization posterior probabilities (CLPP)^7^ with fine-mapping results from GTEx v8^33^ to identify genes where the causal variant is shared between the complex trait and the gene expression trait.
3. We identified the closest gene to the lead variant.

These locus-based methods rely on GWAS data at the locus of interest, thus neither of the evaluation methods described above are applicable. Curated gold standard gene sets are often biased towards well-studied and more readily identifiable causal genes, so we leveraged our fine-mapping data to construct a large set of gold standard genes using fine-mapped protein coding variants. Specifically, we identified 1,348 1 Mb loci centered at non-coding credible sets that additionally contained at least one of a set of 589 gold standard genes harboring a fine-mapped (PIP > 0.5) protein-coding variant for the same trait. This construction formalizes our intuition that an associated protein coding variant is often the relevant functional variant and that a nearby, secondary signal is much more likely to act through this gene than through a separate, distinct gene (see Discussion).

We evaluated the performance of each method in these non-coding loci (after removing coding signals, see Methods) using precision, the fraction of prioritized genes in the gold standard set, and recall, the fraction of gold standard genes in the prioritized set. We first confirm that PoPS outperforms DEPICT and NetWAS using these metrics (**Supplementary Fig. 5, Supplementary Table 4**). Before comparing locus-based methods to each other and to PoPS, we evaluated multiple prioritization criteria using both absolute thresholds and relative rank within a locus for each method (**Supplementary Fig. 6, Supplementary Table 5**). For many methods including TWAS, PCHi-C, and E-P correlation, it is common to prioritize any gene with a predicted connection or significant association; however, across all methods, we found that prioritizing the best ranked gene in the locus had higher precision than including any gene passing a global threshold. These results are consistent with the idea that a regulatory variant can affect the expression of multiple genes^32^, yet only a select few of these genes, perhaps often the most strongly regulated, have a direct effect on the complex trait of interest. Our evaluation of different criteria for each locus-based method led us to prioritize genes that passed global thresholds that were also the best ranked gene in the locus (see Methods).

All individual methods for prioritizing genes from the non-coding signal showed precision less than 50% except CLPP, which had both the highest precision, 52%, and the lowest recall, 4% (**Fig. 4a, Supplementary Table 4**). PoPS and distance performed similarly to each other and had the next highest precision, 47% and 46%, and the highest recall, 47% and 48%, respectively. Other methods, including TWAS, E-P correlation, and ABC-Max, yielded comparable precision but substantially lower recall (10-27%). The low recall of these methods can be attributed in part to limited power to disentangle the causal variant from LD and to missing trait-relevant cell types in the variant-to-gene regulatory maps. These challenges may also contribute to the variability of precision and recall estimates observed across traits (**Supplementary Fig. 7, Supplementary Table 6**). Overall, few genes could be prioritized with confidence greater than 50%, so we sought to devise an approach achieving greater precision.

**Figure 4.**
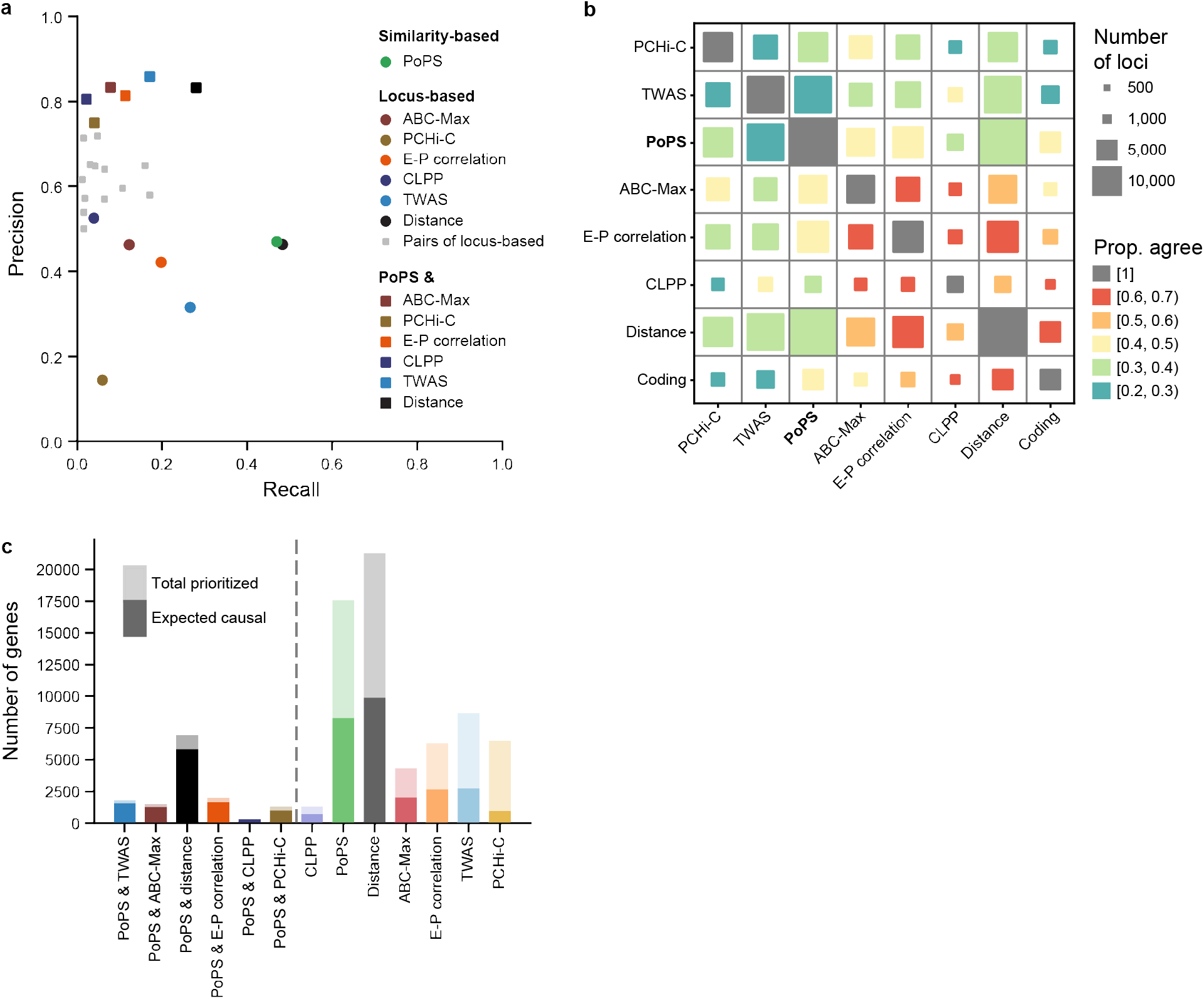
Comparing and combining PoPS with locus-based methods. **a**, Precision-recall plot showing performance of locus-based methods, PoPS, intersections of pairs of locus-based methods, and intersections of PoPs with locus-based methods on the gold-standard set of 589 genes with fine-mapped protein coding variants. Precision is the proportion of prioritized genes in the gold standard set. Recall is the proportion of genes in the gold standard set that were prioritized. **b**, Overlap and agreement among methods across all genome-wide significant loci. Each square represents a pair of methods. The size corresponds to the number of loci where both methods prioritize a gene, and the color corresponds to the proportion of these loci where both methods prioritize the same gene. **c**, Number of unique gene-trait pairs prioritized across all genome-wide significant loci by PoPS, locus-based gene prioritization methods, and intersections of PoPs with locus-based methods, sorted by estimated precision. The full height of each bar represents the total number of genes prioritized. The dark portion of each bar represents the expected number of true causal genes prioritized. Methods to the left of the dashed line achieve precision greater than 75%.

### Combining PoPS with locus-based methods

Towards improving overall gene prioritization, we investigated the extent of agreement among methods evaluated above, additionally comparing to genes with fine-mapped (PIP > 0.1) coding variants. For each pair of methods, we computed the number of loci in which both methods prioritized a gene and the proportion of those loci where they prioritized the same gene (**Fig. 4b, Supplementary Table 7**). Overall, we found low concordance among methods. For example, PCHi-C prioritized a gene in 8,777 loci, while ABC-Max prioritized a gene in 7,913 loci, yet there are only 5,196 loci where both methods prioritized any gene, and of these loci, the two methods agreed only 42% of the time. In general, ABC-Max, E-P correlation, CLPP, and coding agreed more with each other than other methods but prioritized many fewer genes overall. PoPS had mild agreement with most other methods, prioritizing the same gene in up to 43% of loci. Together with the results on gold standard genes, these results suggest that PoPS is a less redundant but similarly accurate method compared to existing locus-based methods and may help to identify a unique set of causal genes.

The range of precision and recall of individual methods and their overall lack of agreement suggests that combining methods may enable better performance for gene prioritization. We evaluated the performance of intersecting pairs of methods, *i.e*. prioritizing genes nominated by both methods individually, on our set of gold standard genes (**Fig. 4a, Supplementary Table 4**). First, we observed that intersecting pairs of locus-based methods often increased precision but never yielded precision above 71%. Second, we observed that intersecting PoPS with any locus-based method yielded precision of at least 75% and up to 86%. Notably, PoPS intersected with TWAS sees the greatest increase in precision and achieves the highest precision of any combination of methods, increasing precision for TWAS from 32% to 86% while maintaining 17% recall. We also find that PoPS intersected with distance increases precision from 46% to 83%, while achieving 28% recall, suggesting that PoPS can substantially improve the precision of the commonly-used nearest gene approach. We conclude that leveraging the polygenic patterns learned by PoPS along with locus-specific genetic information will boost gene prioritization efforts.

### High-confidence prioritized genes

After demonstrating that we can prioritize genes with high precision on our set of gold standard genes, we prioritized genes across all genome-wide significant loci for 95 UK Biobank traits and 18 additional complex traits for which we only had summary statistics. For the UK Biobank traits, we intersected PoPS with TWAS, distance, ABC-Max, E-P correlation, CLPP, or PCHi-C to prioritize between 353 and 6,961 genes each, with estimated precision greater than 75% (**Fig. 4c, Supplementary Table 8**). For the traits for which we only had summary statistics, we intersected PoPS with TWAS or distance, prioritizing 47 and 240 genes, respectively, with estimated precision greater than 83% (**Supplementary Fig. 8, Supplementary Table 9**). In total, we prioritized 8,402 unique gene-trait pairs in 45% of loci with greater than 75% precision.

Genes with high PoP scores and support from multiple lines of evidence include many well-known causal genes (**Fig. 5**). For example, the lipid metabolism genes^34^, *APOE, APOA1, LDLR, APOB*, and *CETP*, were the top 5 genes by PoP score for LDL cholesterol. For mosaic loss of Y (LOY) chromosome in circulating blood^35^, a phenotype with genetic relevance to multiple malignancies, the top genes by PoP score are involved in the DNA damage response (*TP53*) and apoptosis (*BCL2, BAX, BCL2L11*). For schizophrenia, the top genes by PoP score are the well known dopamine receptor (*DRD2*)^36,37^, calcium channel genes (*CACNA1C, CACNB2*)^31^, an important transcriptional regulator underlying developmental delay (*BCL11A*)^37,38^, and the gene encoding for the histone acetyltransferase p300.

**Figure 5.**
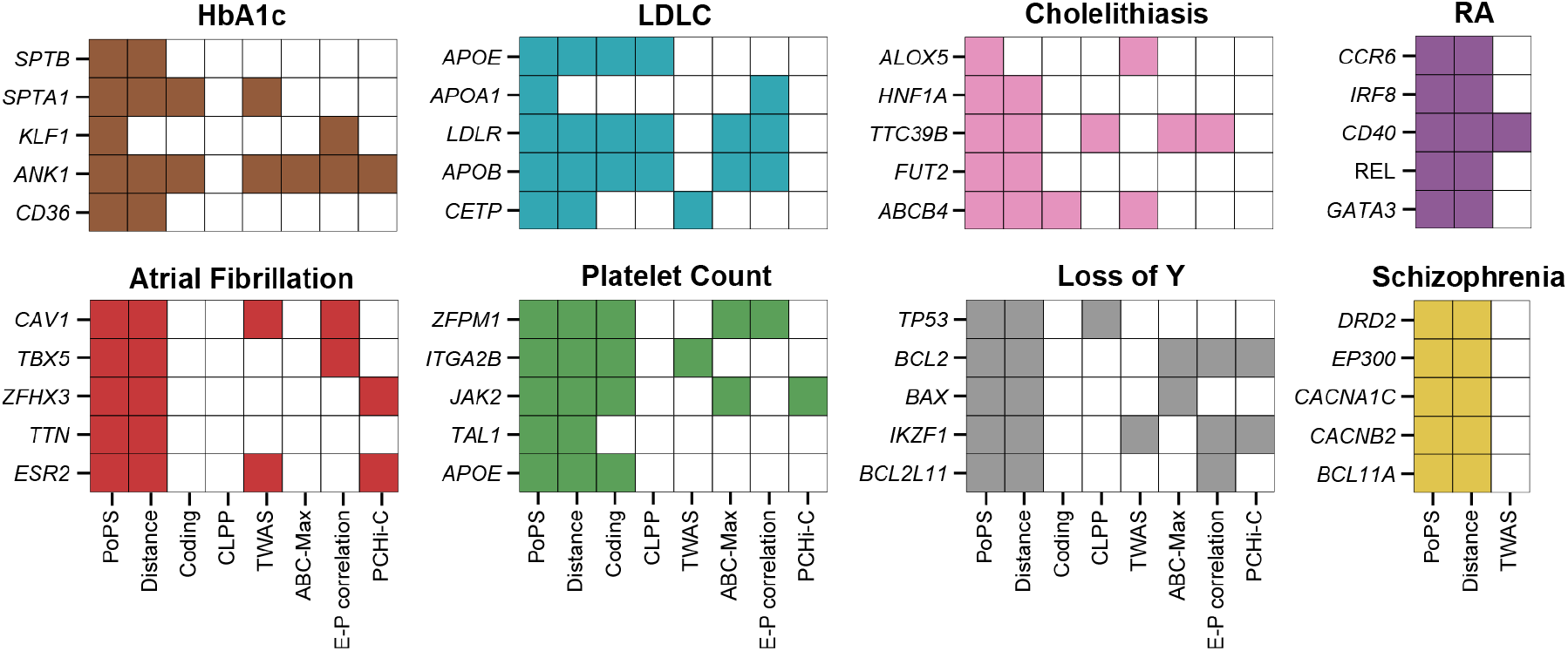
High confidence genes for selected traits. Top five genes prioritized by PoPS and at least one locus-based method, ranked by PoP score, for selected traits. Shaded boxes indicate if a method prioritized the gene.

We highlight four specific examples where the causal variant has been shown to regulate multiple genes or where the causal gene is not the nearest gene, but PoPS, leveraging polygenic patterns, uniquely identified the causal gene. First, for multiple red blood cell traits, rs1175550 was fine-mapped with PIP > 0.9, predicted to be an eQTL for *SMIM1* and *LLRC47* by CLPP, and demonstrated experimentally to affect the expression of *SMIM1, LRRC47*, and *CEP104*^39^. In the locus, PoPS correctly prioritized *SMIM1*, which encodes for the rare Vel blood group protein involved in red blood cell production^40^ (**Fig. 6a**). Also for red blood cell traits, the variant rs737092 was fine-mapped with PIP = 0.72 for mean corpuscular hemoglobin (MCH), and experimental evidence shows that the expression of both *RBM38* and *RAE1* are affected by this variant^39^ (**Fig. 6b**). While other methods do not agree on a causal gene for this locus, PoPS prioritized RBM38, which has been shown to play a role in splicing key erythroid transcripts during terminal erythropoiesis^39^. Next, at the 1p13 locus which is associated with LDL cholesterol and several cardiovascular traits, the variant rs12740374 was fine-mapped with PIP > 0.99, co-localized (CLPP > 0.9 for all) with an eQTL for the proximal genes *CELSR2* and *PSRC1*, as well as *SORT1* (**Fig. 6c**). Here, PoPS is able to correctly identify *SORT1*, which has been shown to encode for a protein that modulates hepatic very low-density lipoprotein levels, altering plasma LDL-C^41^. Finally, for bone mineral density (BMD), the variant rs1550270 falls in an intron of *MSMO1*, was fine-mapped with PIP = 0.39, co-localized with an eQTL for *CPE* in osteoclasts^42^, and was predicted to have a role in regulating *CPE* as well as four other nearby genes (**Fig. 6d**). PoPS correctly prioritized *CPE*, the knockout of which in mice resulted in low bone mineral density and showed indications of increased bone turnover^43^. These examples demonstrate the power of PoPS to provide an important line of independent evidence in cases where the causal variant can be tied to multiple candidate genes and current methods are unable to distinguish the causal gene.

Leveraging PoPS with locus-specific information, we propose a potential novel gene for cholelithiasis (gallstones), along with the causal variant and variant-to-gene mechanism. Gallstones are small stones, often composed of cholesterol, that form in the gallbladder due to excess cholesterol in bile^44^. The variant rs686030:C>A, intronic to *TTC39B*, showed genome-wide significant marginal association and was fine-mapped with PIP = 0.38 for gallstones, although fine-mapping for the pleiotropically associated traits HDL cholesterol and ApoA levels resolved this association down to a single variant (PIP > 0.98 for both, **Fig. 7a**). We found five genes in the locus within 500 Kb of this lead variant but overwhelming support for *TTC39B* from PoPS, ABC-Max, E-P correlation, and CLPP (**Fig. 7b**). A previous pathway analysis failed to find support for *TTC39B* in gallbladder disease^45^ but, here, PoPS uniquely identified *TTC39B* as the causal gene. In the GTEx fine-mapping results, rs686030:C>A demonstrated the greatest effect on reducing the expression of *TTC39B* in the liver compared to other tissues (*β* = −0.31, PIP = 0.43). In other work, decreased TTC39B has then been shown to result in increased levels of HDL cholesterol in mouse knockdown and knockout studies, although the exact tissue in which TTC39B functions to regulate HDL cholesterol levels remains unclear^27,46^. By leveraging our fine-mapping results, we find that rs686030 lies within a relatively tissue-specific regulatory region (accessible in 22/733 cell-types biosamples). Of note, we observed that this region is not only accessible in certain liver and intestinal cell-types, consistent with the results from conditional knockout mice, but that it is most accessible in white blood cell types, such as B cells and monocytes (**Fig. 7c**). Therefore, we hypothesize that rs686030:C>A reduces the expression of *TTC39B* via an enhancer in liver, intestinal, or white blood cell subsets, resulting in increased levels of HDL cholesterol and the formation of gallstones.

**Figure 6.**
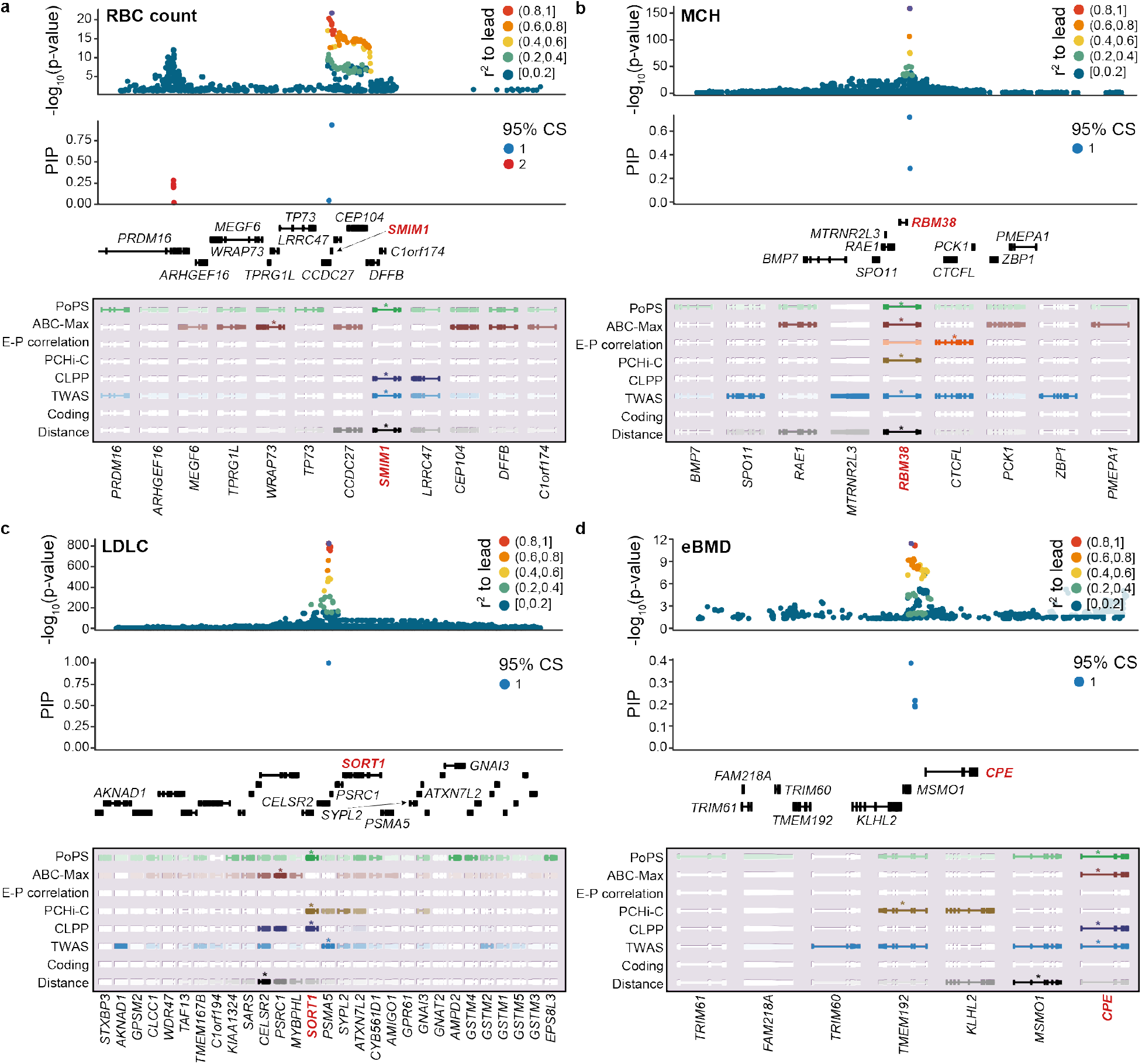
Known biological examples. Top: summary statistics colored by LD to the lead variant and fine-mapping results for variants in the locus colored by credible set. Bottom: results from PoPS and locus-based methods for all genes in the locus. Genes are colored by strength of prediction for each method, with a star denoting the prioritized gene. **a**, rs1175550, *SMIM1* for red blood cell (RBC) count. **b**, rs737092, *RBM38* for mean corpuscular hemoglobin (MHC). **c**, rs12740374, *SORT1* for LDL cholesterol. **d**, rs1550270, *CPE* for bone mineral density.

**Figure 7.**
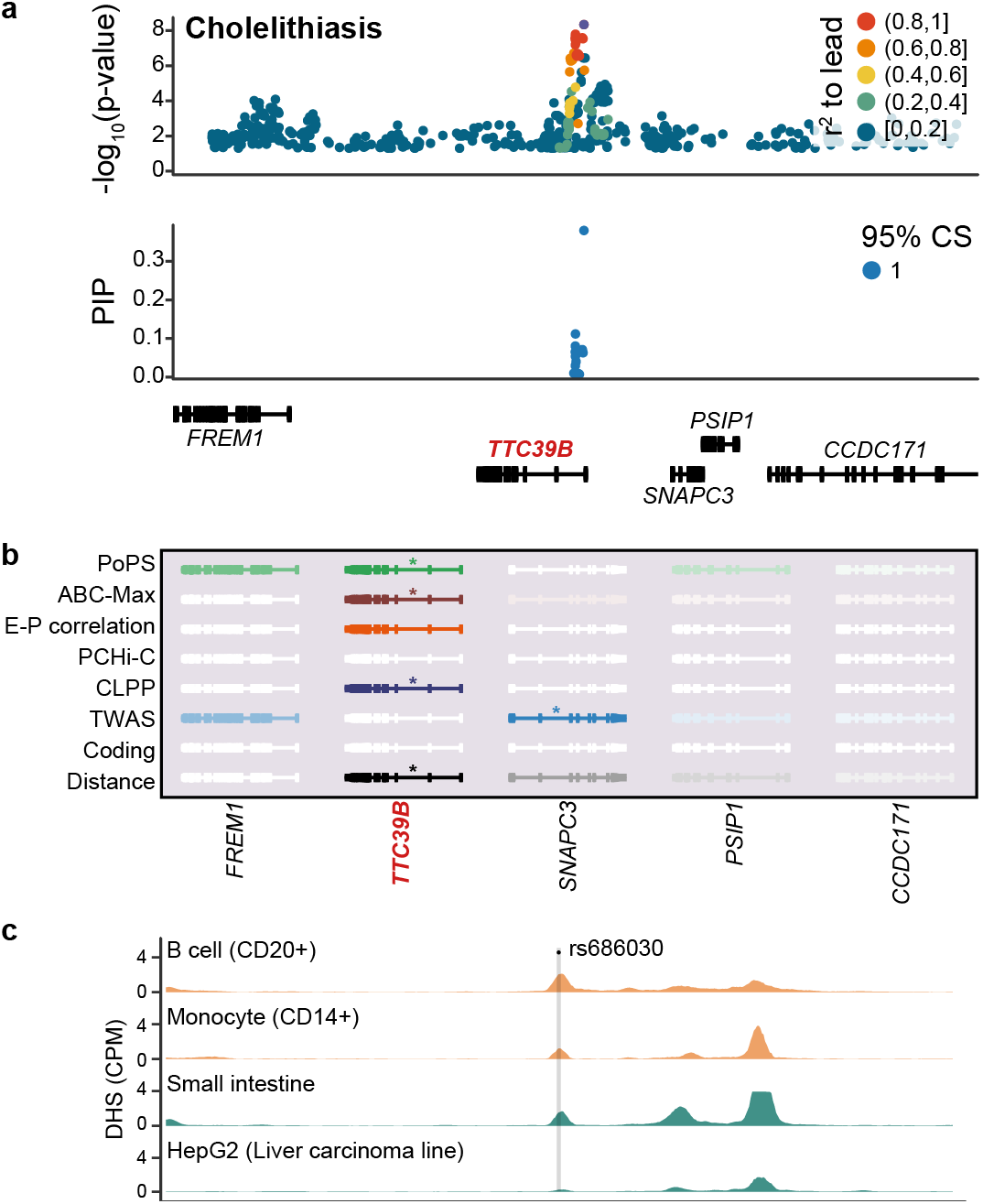
PoPS identifies TTC39B as a candidate gene for Cholelithiasis. **a**, Summary statistics colored by LD to the lead variant, rs686030, and fine-mapping results for variants in the locus colored by credible set. **b**, Results from PoPS and locus-based methods for all genes in the locus. Genes are colored by strength of association for each method, with a star denoting the prioritized gene. **c**, DNase I hypersensitivity tracks shown in read counts per million for four cell-types where rs686030 lies in accessible chromatin.

## DISCUSSION

We developed a new computational method, PoPS, for prioritizing causal genes from GWAS that leverages the genome-wide genetic association information and integrates data from gene expression data sets, protein-protein interaction networks, and pathway databases. We applied PoPS to summary statistics from 113 traits and used unbiased metrics to validate and show that PoPS improves upon other similarity-based methods. In addition, we benchmarked several existing locus-based gene prioritization methods and demonstrated that PoPS performs strongly in comparison while leveraging a distinctive signal. Finally, we proposed a strategy for prioritizing genes more confidently by combining PoPS with existing locus-based methods and achieved precision on a large set of gold standard genes above 75%. To our knowledge, our approach nominates causal genes at non-coding GWAS loci with greater confidence than any existing method. These findings highlight the importance of leveraging data at multiple levels and, specifically, combining gene prioritization results from locus-based and similarity-based methods. Although our polygenic approach to gene prioritization allows for confident prediction of causal genes, it has several limitations. First, our approach relies on the assumption that causal genes share biological characteristics captured by the gene features included in the model. Causal genes that act through unrelated mechanisms or genes whose shared functions are not described by our features would not be identified using PoPS. As new gene expression, protein network, and pathway datasets are generated and incorporated into PoPS, its performance will improve. Furthermore, to leverage the polygenic signal, we assume the causal mechanisms are shared between top loci and sub-significant loci. Second, while informative for ranking genes, the polygenic priority score lacks interpretable units, is not comparable across traits, and does not quantify the uncertainty in the predictions. Third, PoPS alone does not provide a prediction for the causal variant or a variant-to-gene mechanism for prioritized genes. Fourth, PoPS does not directly link causal genes with their relevant cell types. The joint linear model includes many highly correlated features, hindering the interpretability of the features and their coefficients. Although we identified meaningful feature clusters with large contributions to PoP scores for prioritized genes, we leave further investigation of the pathways, cell-types, and tissues predicted by PoPS to future work.

We leveraged genes with fine-mapped coding variants and nearby non-coding signals to evaluate similarity-based methods, locus-based methods, and combinations of methods. This set serves as a powerful tool that, unlike other commonly-used gold standard gene sets such as Mendelian disease genes or drug targets, allows us to evaluate both similarity-based methods that leverage pathway databases and locus-based methods that leverage local signal across a large number of traits and in a framework that is unbiased by previous trait-specific knowledge. However, we note that this set also has several limitations. First, the accuracy of the validation rests on the assumption that non-coding signals near genes with fine-mapped coding variants are explained by the same gene. As the number of independent signals discovered for highly polygenic traits continues to grow, the likelihood that two nearby signals do not point to the same gene also grows; thus, this evaluation may be less accurate for traits with very large numbers of independent associations (**Supplementary Fig. 9**). Second, the set of genes harboring fine-mapped coding variants may skew towards less constrained genes, introducing a bias. Finally, many methods rely on having data measured in the specific cell-types and tissues relevant to the trait of interest. In the evaluation framework described here, it is not possible to distinguish whether the performance of a method is limited by the methodology or the availability of the necessary data.

In conclusion, leveraging polygenicity improves causal gene prioritization. In doing so, PoPS is a powerful tool for identifying causal genes from GWAS summary statistics and marks an important step towards building functional understanding from genetic associations. The ability to more confidently prioritize causal genes will aid in understanding the underlying trait biology and nominate genes that are strong candidates for experimental follow-up.

### URLs

Gene features, https://github.com/FinucaneLab/gene_features; fine-mapping in the UK Biobank, https://www.finucanelab.org/data; precomputed TWAS weights for GTEx v7 tissues, http://gusevlab.org/projects/fusion/weights/GTEX7.txt.

### Data availability

A repository of processed gene features, visualizations of top derived features, and code to reproduce these analyses are available on GitHub at https://github.com/FinucaneLab/gene_features. Complete PoPS results for 95 complex traits in the UK Biobank and 18 additional disease traits as well as results for PoPS and locus-based methods in genome-wide significant loci are available at https://www.finucanelab.org/data.

### Code availability

PoPS is available as an open-source Python package at https://github.com/FinucaneLab/pops.

## Acknowledgements

We thank Krishna Aragam, Adam Butterworth, Mark Daly, Nikita Artomov, Yakir Reshef, and all members of the Finucane lab for helpful discussions. This research was conducted using the UK Biobank Resource under project 31063. H.K.F. was funded by NIH grant DP5 OD024582 and by Eric and Wendy Schmidt. J.M.E. was supported by a Pathway to Independence Award (K99HG00917 and R00HG009917), the Harvard Society of Fellows, and the Base Research Initiative at Stanford University. J.M. and J.N.H. were supported by NIH grant R01DK075787. R.S.F. was supported by NHGRI NIH F31HG009850. J.O.M was supported by the Richard and Susan Smith Family Foundation, the HHMI Damon Runyon Cancer Research Foundation Fellowship (DRG-2274-16), the AGA Research Foundation’s AGA-Takeda Pharmaceuticals Research Scholar Award in IBD – AGA2020-13-01, the HDDC Pilot and Feasibility P30 DK034854, and the Food Allergy Science Initiative.

## Competing interests

J.C.U reports compensation from consulting services with Goldfinch Bio and AVROBIO. R.S.F. is an employee of Vertex Pharmaceuticals. C.P.F. is an employee of Bristol Myers Squibb. J.O.M. reports compensation for consulting services with Cellarity. A.R. is a co-founder and equity holder of Celsius Therapeutics, an equity holder in Immunitas, and was an SAB member of ThermoFisher Scientific, Syros Pharmaceuticals, Neogene Therapeutics and Asimov until July 31, 2020. From August 1, 2020, A.R. is an employee of Genentech. J.N.H. served on the Scientific Advisory Board of and consults for Camp4 Therapeutics. E.S.L. serves on the Board of Directors for Codiak BioSciences and Neon Therapeutics, and serves on the Scientific Advisory Board of F-Prime Capital Partners and Third Rock Ventures; he is also affiliated with several non-profit organizations including serving on the Board of Directors of the Innocence Project, Count Me In, and Biden Cancer Initiative, and the Board of Trustees for the Parker Institute for Cancer Immunotherapy. He has served and continues to serve on various federal advisory committees.

## ONLINE METHODS

### MAGMA gene z-scores

We applied MAGMA^9^ to the summary statistics for each trait using the 1000 Genomes Project reference panel^20^ to compute gene-level association statistics and gene-gene correlations using the SNP-wise mean gene analysis and a 0 Kb window around the gene body for mapping SNPs to genes. For each gene, MAGMA computes a gene p-value from the mean chi-square statistic of SNPs in the gene body and its approximate sampling distribution. The gene p-value is converted to a z-score using the probit function. The resulting z-score reflects the gene-trait association after correcting for linkage disequilibrium (LD) among SNPs within the gene body. MAGMA approximates the gene-gene correlation matrix, *R*, using the correlations between the model sum of squares of each gene pair under the joint null hypothesis of no association. These correlations are a function of the LD between SNPs in each pair of genes and represent the LD on a gene level. To ensure a positive semidefinite correlation matrix we add a small value to the entries of *R* along the diagonal. Specifically, if the minimum eigenvalue, *λ*_*min*_, of *R* is less than 0, we add |*λ*_*min*_| + 0.05 to each element along the diagonal.

### PoPS covariates

We included covariates corresponding to gene density, effective gene size, and inverse of the mean minor allele count (MAC) of SNPs in the gene as well as the log of these variables as computed by the MAGMA^9^ software. MAGMA defines gene density as the ratio of the effective number of independent SNPs in the gene to the total number of SNPs in the gene and defines effective gene size as the effective number of independent SNPs in the gene. We additionally include a covariate corresponding to gene size and the log of this variable, defined as the length of the gene in base pairs.

### Locus definition

From the set of associated variants with P < 5×10^−8^, we designated independent lead variants from which to define loci. For the 18 traits where we used publicly available summary statistics, we performed PLINK clumping using EUR individuals in the 1000 Genomes Project reference panel with a p-value threshold of 5×10^−8^ and r^2^ threshold of 0.1. Within each clump, we defined the variant with the most significant p-value as the lead variant. For the 95 traits from the UK Biobank where we had fine-mapping results for regions containing genome-wide significant variants, we defined one locus for every independent credible set (CS). For each fine-mapped CS, we defined the variant with the highest posterior inclusion probability (PIP) as the lead variant. This yields better resolution on the identity of the likely causal variant and allows us to account for multiple independent signals contributing to a single GWAS hit. We then defined the locus boundaries as 500 Kb on either side of the lead variant and included all genes that fell within or overlapped with the locus boundaries. (See **Supplementary Fig. 9** for sensitivity to boundary size.)

### Complex traits and disease associations

GWAS for 95 heritable traits in the UK Biobank were performed as part of an ongoing study (Ulirsch and Kanai, *In preparation*). Up to 361,194 individuals of white British ancestry with available phenotypes and variants with INFO > 0.8, minor allele frequency (MAF) > 0.01%, and Hardy-Weinberg equilibrium (HWE) p-value > 1e-10 were included in the GWAS. Covariates for the top 20 PCs, sex, age, age^2^, sex*age, sex*age^2^, and dilution factor where applicable were controlled for in the association studies. Quantitative traits were inverse rank transformed and associations were estimated using BOLT-LMM^47^. For binary traits, associations were estimated using SAIGE^48^. Publicly available summary statistics were downloaded for an additional 18 diseases (**Supplementary Table 1**).

### Gene features

We created gene features from three main data types: (**1**) bulk and single-cell gene expression datasets, (**2**) curated biological pathways, and (**3**) predicted protein-protein interaction networks.

(**1**) For each of the 77 gene expression datasets (**Supplementary Table 2**), we uniformly reprocessed the raw count (or normalized count when raw counts were not provided) matrices using Seurat v3^49^. First, cells with total counts outside of the 5-95th quantiles were removed and only the 18,383 protein coding genes used in the PoPS analysis were included. Counts were then scaled to counts per million (CPM), log normalized, and scaled such that each gene had a mean of 0 and variance of 1 across cells. Principal components and gene loadings were computed on scaled expression values for the top 1,000-3,000 variable genes using truncated SVD^50^. Independent components and gene loadings were computed using fastICA^51^. A k-nearest neighbor graph was created using the top principal components (PCs, based upon inspection of elbow plot) and clusters were identified using the Louvain algorithm. The uniform manifold approximation and projection (UMAP) algorithm^52^ was used to visualize clusters and investigate batch effects. When batch effects were visually apparent and pre-defined batch annotations were provided, we attempted to remove batch effects using the anchor approach outlined in Stuart et al. Finally, we performed differential expression between clusters using a one-vs-all approach with a two-sided Welch’s *t*-test. We provide code to reproduce these analyses, a repository of processed features, and visualizations of top derived features at https://github.com/FinucaneLab/gene_features.

We then derived features for PoPS (**a**) on the whole dataset, (**b**) within clusters representing different cell populations, and (**c**) between clusters. (**a**) On the whole dataset, we derived features of gene loadings from principal component analysis (PCA) and gene loadings from independent component analysis (ICA). (**b**) Within each cluster, either predefined (when available) or identified in our analysis, we derived features of average scaled gene expression and gene loadings from the top 10 PCs. (**c**) Comparing across clusters (1-vs-all), we derived features of a *t*-statistic for differential expression and a binary indicator for differentially up- and down-regulated genes (Benjamini–Hochberg FDR < 0.05 and |log_2_(fold-change)| > 2).

(**2**) We created features from biological pathways curated for DEPICT from KEGG^23^, Gene Ontology^22^, Reactome^24^, and the Mouse Genome databases^25^. Each feature was encoded as a binary indicator for membership to a pathway. (**3**) We created features using the predicted InWeb_IM protein-protein interaction (PPI) network^21^. For each gene, we included as a feature a binary indicator for the set of genes that were its first-degree neighbors.

Finally, for each distinct dataset, we included a control feature as a binary indicator for the set of genes that were reported in that dataset. All features were centered and scaled to have mean of 0 and variance of 1 across genes.

### Benchmarker

Benchmarker^26^ is an unbiased, data-driven approach to evaluate gene prioritization methods. Based on the assumption that SNPs near causal genes should be enriched for trait heritability, Benchmarker uses stratified LD score regression (S-LDSC)^53^ to estimate the average contribution of SNPs near prioritized genes to per SNP heritability. Using S-LDSC, Benchmarker jointly models a SNP annotation corresponding to prioritized genes along with 53 annotations in the “baseline model” which include genic, regulatory, and conserved regions. To evaluate performance, we use the regression coefficient, **τ**, and its p-value for the hypothesis **τ** > 0. **τ** measures the contribution of SNPs near prioritized genes to per SNP heritability after controlling for the baseline annotations. To make **τ** comparable across traits, we normalized **τ** by the average per-SNP heritability for each trait and refer to this quantity as normalized **τ**.

For our analyses, we selected the 500 genes with the highest PoP scores for each trait as the set of prioritized genes and used a 100 Kb window on either side of the transcription start site of each gene for mapping SNPs to genes.

### Closest gene enrichment

We used a normal approximation to the null distribution for our test statistic, *c*, the number of genes that are PoPS prioritized and the closest gene to the lead variant in a locus. Under the null, PoPS prioritizes the closest gene in a locus at random with probability 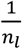, where *n*_*l*_ is the number of genes in a locus, *l*. Across all *L* loci, the distribution of *c* under this null is a sum of independent Bernoullis with different biases. For computational tractability when *L* is large, we approximate this by a normal with matched moments.

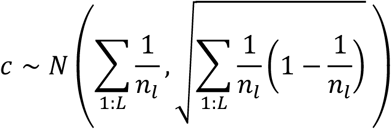

We performed a one-sided test for 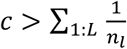 under the null. We additionally computed the enrichment of the number of PoPS prioritized genes that are the closest as the ratio of the observed to the expected, 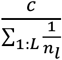, and estimated the standard error of the enrichment. We used the bootstrap to estimate the standard error of the enrichments, not assuming a null distribution, and performed 1024 bootstrap repetitions resampling the *L* loci for each trait.

### Independent traits

To identify independent traits, we first computed genetic correlations between all pairs of traits using cross-trait LD score regression^54^ with LD scores from UK10K^55^. Next, we created an adjacency matrix of traits with edge weights corresponding to whitened (|r_g_| < 0.2 were set to 0), absolute genetic correlations. We then identified the maximum independent set of vertices (traits) such that no two were adjacent using the igraph package^56^ in R 3.5. The resulting set contained 46 independent traits (**Supplementary Table 1**).

### Feature clustering

For each trait, 50 PCs were derived from the scaled gene by feature matrix using truncated SVD. A feature by feature distance matrix was then created as the dissimilarity between features using one minus the squared Pearson correlation (*r*^2^) between PCs. Complete linkage hierarchical clustering was then performed on this distance matrix. Clusters were determined such that Pearson *r*^2^ > 0.1^2^ for all features within a cluster. This inclusive threshold was chosen in order to reduce the impact of multicollinearity when interpreting the contribution of top clusters to PoP scores and was validated by manual investigation of within-dataset composition of large clusters as well as biological interpretability of the top clusters.

### Fine-mapping

Fine-mapping aims to disentangle the effects of LD to identify causal genetic variants underlying complex trait associations and was performed as part of an ongoing study (Ulirsch and Kanai, *In preparation*). Fine-mapping was performed using the Sum of Single Effects (SuSiE) method^57^, allowing for up to 10 causal variants in each region. Prior variance and residual variance were estimated using the default options, and single effects (potential 95% CSs) were pruned using the standard purity filter such that no pair of variants in a CS could have r^2^ > 0.25. Regions were defined for each trait as +/− 1.5 Mb around the most significantly associated variant, and overlapping regions were merged. As inputs to SuSiE, summary statistics for each region were obtained using BOLT-LMM^47^ for quantitative traits and SAIGE^48^ for binary traits, in sample dosage LD was computed using LDStore^58^, and phenotypic variance was computed empirically. Variants in the MHC region (chr6: 25-36 Mb) were excluded as were 95% CSs containing variants with fewer than 100 minor allele counts. Coding (missense and predicted loss of function) variants were annotated using the Variant Effect Predictor (VEP) version 85^59^.

### ABC-Max

We used the Activity-by-Contact (ABC) model^11^ to predict enhancer-gene connections in 131 biosamples from 74 distinct cell-types and tissues based on measurements of chromatin accessibility (ATAC-seq or DNase-seq) and histone modifications (H3K27ac ChIP-seq). For each trait, we included only predicted enhancer-gene connections where the enhancer contained a fine-mapped variant (PIP > 0.1) in a credible set that did not contain any coding or splice site variants. We assigned each gene in a locus a single score for the corresponding fine-mapped CS by taking the highest ABC score of predicted enhancers for that gene-CS pair across all biosamples that are enriched for overlapping fine-mapped variants for that trait. Finally, to predict a single gene for each credible set, ABC-Max prioritizes the gene with the highest ABC score in the locus.

### Enhancer-promoter correlation

We downloaded predicted enhancer-promoter maps based upon the correlation of biochemical marks at regulatory regions and expression of nearby genes across cell types for 808 tissues and cell-lines from the FANTOM5 project^15^, 127 tissues and cell-lines from the ROADMAP Epigenomics project^16^, and also for 16 primary blood cell types^13^. For the FANTOM5 dataset, we filtered to interactions with Benjamini–Hochberg FDR < 10^−5^ for a non-zero Pearson correlation. For the ROADMAP dataset, we filtered to interactions with a confidence score > 2.5. For the Ulirsch et al. dataset, we filtered to interactions with a Pearson correlation > 0.7 and a Storey FDR < 10^−4^. Finally, for each trait, we included only predicted interactions where the enhancer contained a fine-mapped variant (PIP > 0.1). We assigned each gene in a locus a single score for each corresponding fine-mapped CS by taking the highest confidence score or correlation of predicted enhancers for that gene-CS pair across all tissues and cell-lines.

### Promoter capture Hi-C

We downloaded promoter capture Hi-C datasets (PCHi-C) containing observed physical interactions between fragmented DNA and targeted genic promoters for 28 diverse human tissues and cell-lines^12^ and 15 primary blood cell types^14^. For the Jung et al. dataset, we filtered to interactions with p-values for interaction < 0.01 and raw frequency counts > 5. For the Javierre et al. dataset, we filtered to interactions with CHiCAGO^60^ scores > 5. In both cases, we defined a variant-gene interaction as a variant with PIP > 0.10 overlapping with a relevant region of accessible chromatin, based upon 39 ROADMAP tissues^61^ for Jung et al. and 44 primary blood cell types^13,62^ for Javierre et al.). Finally, for each trait, we included only predicted interactions where the enhancer contained a fine-mapped variant (PIP > 0.1). We assigned each gene in a locus a single score for each corresponding fine-mapped CS by taking the highest connection strength of predicted enhancers for that gene-CS pair across all tissues and cell-lines.

### TWAS

We applied TWAS^8^ using the FUSION software package and precomputed expression reference weights for 48 tissues from GTEx v7^32^. To avoid leveraging the coding signal for the precision-recall analysis, we excluded all variants in LD (r^2^ greater than 0.2 to a coding variant with PIP > 0.1). For all other analyses we included all variants in the GWAS summary statistics. In both cases, we took the most significant association across tissues for each gene.

### Co-localization

Using fine-mapping results for 95 complex traits from the UK Biobank and for eQTLs in 49 tissues from GTEx v8^33^ we computed co-localization posterior probabilities (CLPP), analogous to those reported by the eCAVIAR software^7^. For each variant, *i*, fine-mapped for a complex trait, *g*, and an eQTL trait, *e*, the CLPP was computed as *P*(*C*_*ig*_, *C*_*ie*_) = *P*(*C*_*ig*_)*P*(*C*_*ie*_), where *P*(*C*_*ig*_) is the PIP of variant *i* in complex trait *g* and *P*(*C*_*ie*_) is the PIP of variant *i* in eQTL trait *e*. This quantity is an estimate of the probability that the variant is causal for both the complex trait and the gene expression trait. Within each fine-mapped CS and for each gene, we took the maximum CLPP across all variants and GTEx tissues.

### Gene prioritization criteria

We evaluated multiple prioritization criteria for each locus-based method and PoPS including various absolute thresholds and the relative rank of genes within a locus (**Supplementary Fig. 6, Supplementary Table 5**). We chose the following prioritization criteria to maximize precision:

(**1a-c**) E-P correlation, PCHi-C, ABC-Max: for each locus such that at least one gene has a predicted connection with an enhancer containing a variant with PIP > 0.1, the gene that has the highest correlation or connection score. To combine across datasets for E-P correlation and PCHi-C, we included any gene prioritized in at least one dataset.
(**2a**) TWAS: for each locus such that at least one gene is significantly associated after Bonferroni correction, the gene with the most significant p-value.
(**2b**) CLPP: for each locus such that at least one gene has a variant with CLPP > 0.1, the gene with the highest CLPP.
(**3**) Distance: for each locus, the gene that is closest to the lead variant by distance to the gene body.
(**4**) PoPS: for each locus, the gene that has the highest PoP score.

### DNase I hypersensitivity annotations

We lifted over uniformly re-processed DNase I hypersensitivity data^63^ for 733 biosamples from GRCh38 to hg19 to investigate the potential cell-type of action for fine-mapped causal variants. Data are shown in aligned read counts per million for ENCODE samples ENCFF220IWU, ENCFF619LIB, ENCFF659BVQ, and ENCFF842XRQ.

